# Barriers to reaching the Zero-Dose and Under Immunized Children in Uganda: a qualitative rapid assessment

**DOI:** 10.1101/2025.10.20.25338036

**Authors:** Emmanuel Mugisha, Shadiah Nanteza Mugizi, Deogratias Agaba, Jacqueline Anena, Joanita I. Nankabirwa, Susan Nayiga, Carol Kamya, Faith Namugaya, Paul Katamba, Patrick Albert Ipola, Fred Isaasi, Peter Waiswa, Allen Kabagenyi, Miriam Kayendeke, David Wafula, Betty Mirembe, Moses R. Kamya, Mayora Chrispus

**Affiliations:** PATH Country Program, Kampala, Uganda; Infectious Diseases Research Collaboration, Kampala, Uganda, School of Medicine, Makerere University Kampala, Kampala, Uganda; Infectious Diseases Research Collaboration, Kampala, Uganda; Makerere University School of Public Health, Kampala, Uganda; Makerere University School of Statistics and Planning, Kampala, Uganda

**Keywords:** Zero Dose, Under-immunized, Vaccination, immunization, barriers, coverage, equity, Uganda

## Abstract

**Background:** Despite great strides made in improving immunization coverage in Uganda, the burden of under-immunized (UI) and zero-dose (ZD) children remains high, increasing the risk of Vaccine Preventable Disease outbreaks. This study explored the barriers to reaching ZD and UI children in selected high-burden communities in Uganda.

**Methods:** This cross sectional qualitative rapid assessment study was conducted in three high-burden districts—Kasese, Mubende, and Wakiso in Uganda The study received ethical approval, and all participants were consented before interview. Data were collected through key informant interviews with local leaders, district health team members, health workers and village health team members, as well as in-depth interviews with caregivers of children aged 18 weeks to 23 months. The collected data were analyzed thematically with the aid of NVIVO 14 (QSR International). Results are presented in themes and subthemes and supported by verbatim quotations where necessary.

**Results:** The study identified a range of demand and supply-side barriers hindering access to immunization for ZD and UI children in Uganda. These barriers are reported under seven themes namely: limited physical access to immunization services; inadequate client-centered immunization services (Long waiting time at health facilities, poor health worker attitudes, immunization-related costs); vaccine stockouts; fear of adverse events following Immunization; limited spousal support; myths and misconceptions related to immunization; and persistent home-based births or deliveries.

**Conclusion:** Understanding the barriers to immunization uptake among zero-dose and under-immunized children is a critical foundation for strengthening vaccination coverage and reducing the incidence of VPDs in Uganda. This rapid assessment conducted in Kasese, Mubende, and Wakiso districts highlights both demand and supply side challenges that hinder access and utilization of immunization services. Addressing these barriers requires coordinated, multi-sectoral approaches that are both comprehensive and contextually grounded.

## Introduction

Vaccine Preventable Diseases (VPDs) remain a significant contributor to the burden of child mortality and morbidity in Sub-Saharan Africa (SSA), with an associated 2.4 million deaths among children annually. It is estimated that 14.3 million children remained unvaccinated (zero-dose children) and 5.6 million were partially vaccinated globally in 2024, with most of these children residing in SSA [1]. In Uganda, immunization coverage has improved overtime with most antigens in the routine immunization schedule registering more than 90% coverage [2]. This improved coverage has been attributed to the accelerated efforts of the Uganda National Expanded Program on Immunization (UNEPI) through implementation of various vaccination strategies including community-based outreaches, home-based vaccination, Integrated Child Health Days (ICHD), mass immunization campaigns, and the Reach Every Child (REC)/ Reach Every District (RED) strategy among others [3].

Despite these efforts, a significant number of zero-dose (ZD) and under-immunized (UI) children still exist in Uganda, contributing to persistent outbreaks of VPDs. According to data from the District Health Information System-2 (DHIS2), a total of 109,338 children did not receive the first dose of Diphtheria-Pertussis-Tetanus (DPT), while 313,467 did not receive DPT3 in 2023 [4]. There are also notable variations in immunization coverage across districts, as well as in rural and urban settings in Uganda [5].

Various studies and assessments have identified several demand and supply drivers for vaccination coverage and uptake in Uganda. These include vaccine stocks; health worker attitudes; community knowledge, attitudes, and perceptions about vaccination; education of caregivers; household socioeconomic status; health-seeking behaviour of households and physical location, among others [4, 6, 7]. While this evidence exists, the burden of ZD children and UI children continues to be disproportionately high in some districts [8]. Reaching these ZD and UI children with life-saving vaccines requires context-specific data and evidence about who they are, where they live, and why they remain unvaccinated [7].

Gavi supports the Zero-Dose Learning Hubs (ZDLH) in Bangladesh, Mali, Nigeria, and Uganda as a key initiative to learn and address immunization equity by identifying and reaching ZD and UI children using high-quality data. As part of the Uganda ZDLH implementation, we conducted a qualitative rapid assessment to identify the barriers and gaps that contribute to low uptake and or completion of immunization schedules in three high burden districts (Kasese, Mubende, and Wakiso). This assessment was to generate evidence to inform the design and implementation of evidence-based and targeted interventions that improve immunization coverage and reduce the burden of ZD and UI children in Uganda.

## Methodology

### Study design and participants

This was a cross sectional qualitative rapid assessment study conducted between 1^st^ August 2023 and 30^th^ March 2024 using key informant interviews (n=24) with Village Health Team (VHT) members, local community leaders and health workers; and in-depth interviews (n=37) with primary caregivers of ZD and UI children aged 18 weeks to 23 months (**Table 1**)

**Table 1:** Interviews conducted under the rapid assessment.

| Respondent | Mubende | Kasese | Wakiso | Total |
| --- | --- | --- | --- | --- |
| VHTs | 3 | 4 | 2 | 9 |
| Caregivers | 12 | 11 | 14 | 37 |
| Local Leaders | 2 | 2 | 2 | 6 |
| Health Workers | 1 | 5 | 3 | 9 |

A zero-dose child was defined as a child aged between 12 months and 23 months who had not received DPT1, and an under-immunized child was defined as a child aged between 4.5 and 23 months who had not received DPT3 at the time of the study (1). Respondents were purposively selected across the three study districts, and interviews were conducted until topical and data saturation was reached.

### Study area and setting

This rapid assessment was conducted in Uganda’s Zero-Dose Learning Hub (ZDLH) districts—Kasese, Mubende, and Wakiso. The districts were purposively selected for their high burden of ZD children and low DPT1 coverage, as reported in DHIS2 data from August 2023 to March 2024. These districts also include communities with equity reference groups (ERGs), reflecting significant immunization inequities. As part of the Equity Accelerator Fund (EAF) initiative, the ZDLH aims to understand the ZD challenge better and co-design targeted interventions to identify and reach these children. A detailed profile of the selected districts is available elsewhere [9]. Sub-counties in each district were classified into four categories using DHIS2 data and the Reach Every District/ Reach Every Child (RED/ REC) categorization, with Category 4 indicating poor immunization access and utilization. High zero-dose burden sub-counties within Category 4 were prioritized for the rapid assessment. Through consultations with District and VHTs, high-risk communities were selected, including i) urban communities (Kasokoso and Kiganda villages in Namugongo sub-county, Wakiso district); ii) island/fishing community (Gulwe and Kyanjazi villages in Bussi sub-county, Wakiso district); iii) community at the national border (Kamukumbi village in Isango sub-county, Kasese district); iv) mountainous areas (Bikunya village in Karambi sub-county, Kasese district); and v) underserved communities (Kiranduzi, Lugalama and Bujaala villages in Kiruuma sub-county, Mubende district). Guided by VHTs, the study team identified households with ZD and UI children, supplementing recruitment via snowball sampling for data collection.

### Data collection

To explore barriers to immunization service utilization, topic guides for key informant interviews (KIIs) and in-depth interviews (IDIs) were developed and administered among stakeholders, including health workers, local leaders, and caregivers of ZD and UI children. All interviewing was done through face to face interviews either at home or at workplace. Each respondent was assigned an interview code for reference and no names were recorded to ensure data anonymity. The interviews were conducted by three trained researchers (FI, DW, SNM, 2 males, 1 Female), all experienced in qualitative methods and fluent in English and the predominant local languages— Lhukonzo in Kasese, and Luganda in Mubende and Wakiso. The data collectors had a minimum of bachelor’s degrees and were part of the study team. Health worker interviews were conducted in English and took between 45 minutes to 1 hour. Caregiver interviews were held in community-agreed locations to ensure comfort and privacy. Interview summaries were compiled after each session and reviewed by the research team to identify emerging areas for further exploration. The data collection process was supervised by three senior researchers (EM, JIN, SN, 1 male, 2 female), who held regular debrief meetings with the field team to monitor data quality and thematic saturation. Data collection was concluded once consensus was reached that no new insights were emerging. No refusal for interview was registered.

### Data management and analysis

All interviews were audio-recorded and supplemented with field notes. Transcriptions were completed in English using meaning-based translation by the data collectors, with support from an additional researcher (MK, Female). Transcripts were cleaned and imported into NVivo 14 for thematic analysis using an inductive approach. Eight researchers (SN, PK, PIA, MK, CK, FI, DW, and FN, 4 male, 4 female) with expertise in qualitative research reviewed the transcripts, generated initial codes, and engaged in researcher triangulation to refine and group codes into sub-themes [10]. These sub-themes were then synthesized into broader themes using a deductive framework, culminating in grand narratives that captured participants’ perspectives on barriers to vaccination uptake among ZD and UI children. Anonymized quotes were selected to illustrate key findings and only reference made to the category of respondent while reporting. Meetings of the entire research study team were held to finalize the analytical framework and validate emerging insights. A completed COREQ checklist is included in the supplementary materials to support transparency and rigor in reporting.

### Ethical approval and consent to participate

This study received ethical approval from Makerere University School of Public Health Research and Ethics Committee (SPH-2023-428). The study was also registered by the Uganda National Council for Science and Technology (HS3011ES). Permission to conduct the study were obtained from local government authorities in Kasese, Mubende, and Wakiso districts.

Prior to the interview process, written informed consent was obtained from all participants using the appropriate language they desired and understood. The research associate took time to explain the details, confirmed understanding by asking a few questions to ensure understanding, asking if the respondent had any questions and addressing any concerns raised. After fully understanding and agreeing to participate, the participant then provided a signature or a thumbprint to mark voluntary consent. Additionally, the participants’ rights and autonomy were respected throughout the interviewing process.

## Results

The data revealed two grand narratives of barriers: demand-side-related barriers and supply-side-related barriers. Within these, seven themes emerged including; limited physical access to immunization services; inadequate client-centered immunization services (Long waiting time at health facilities, poor health worker attitudes, immunization-related costs); vaccine stockouts; fear of adverse events following Immunization; limited spousal support; myths and misconceptions related to immunization; and persistent home-based births or deliveries.

### Supply-side related barriers

#### 1. Limited physical access to immunization services

Participants in the In-depth interviews (caregivers) reported challenges related to physical access to immunization services, and this was mainly associated with long distances to health facilities, and fewer and irregular immunization outreach services.

### Long distances and high transport costs to health facilities

Most caregivers reported walking long distances to health centers to seek health care services. Communities reside far from health facilities or service points requiring significant travel time. This is compounded by challenging physical terrains characterized by hills, rivers, swamps, and other natural barriers that complicate movement. While caregivers may sometimes opt to use *bodabodas* (motorcycle taxis), they are limited in these areas and very costly, making them inaccessible for many community members. Sometimes the *bodabodas* are unavailable during adverse weather or at night. It was noted that even after travelling the distances, sometimes, they do not find immunization services and are referred to other health facilities. At some health facilities, health workers sometimes referred caregivers to higher-level health facilities for immunization services, some of which are more than fifteen kilometers away. These factors collectively hinder access to immunization services, particularly for vulnerable populations, like one respondent noted;

> *“When I went to the Health Centre, I found that there was no vaccine, health workers advised me to go to another Health Centre, and when I reached there, there were no vaccines either, and they also advised me to go to the Hospital. The only means to go to the Hospital is by motorcycle and you can find that sometimes we don’t have transport [money] at home, and we end up missing that day of immunization*.*”* ***— IDI, Caregiver, Kasese district***.

In Bussi Island (Wakiso District), participants reported that the health facility was approximately 10 kilometres from the island, which limited their access to immunization services at the facility. They reported that they mostly relied on immunization outreaches. Caregivers in Mubende district reported similar experiences.

> *“There are poor roads, and you find that the health facility where they immunize is very far, which put them off. Another thing is that the money they have could be very little, and they don’t even have 1,000 shillings [30 cents] to buy essentials, leave alone spend this on transportation. They are charged between 5,000 shillings [1*.*4 USD] and 7,000 shillings [2 USD] for transport to take the child for immunization*.**” KII, Community leader, Mubende district**.

> *“I would walk on foot to go to Kitanda Health Centre* [private for-profit clinic]. *It is very far from here and you pass Kiranduzi and go up the hill, yet you have put the child on your back. The mode of transport is hard; you walk a lot and reach when you are tired. It is a long distance*”. **IDI, Caregiver, Mubende district**.

### Few and irregular immunization outreach services

Some participants reported fewer and irregular immunization outreach services, which would ideally provide a substitute for the health facilities that are far from where they live. In Bussi, Wakiso district, where caregivers commonly mentioned that their outreach posts organized within their communities were very helpful in bringing the services closer, there were concerns that the outreaches had since ceased, making it hard for their children to get vaccinated.

*“He [child] was given some vaccines as part of the outreach, but health workers do not come regularly. He stopped being immunized at one and a half [years]”*. ***IDI, Caregiver, Wakiso district***.

The service providers equally appreciated the challenges of immunization due to distance and the high costs that caregivers must incur to access the health facilities. Health workers confirmed that while outreaches helped to extend the services closer to communities and were key in increasing utilization of immunization services, they were irregular due to resource constraints.

> *“We can organize outreaches when a village is very far from the health centre. We are given financial facilitation, but it comes after 3 months or on a quarterly basis, and you can find that the health worker doesn’t have transport to go to that village. You find that children have missed immunization because we have not gone there*.*”* ***KII, Health Worker, Wakiso district***.

#### 2. Inadequate client-centered services

Caregivers elicited three aspects that characterized inadequate client-centered services and constrained utilization of immunization services. These included long waiting hours at health facilities, poor health worker attitudes, and costs associated with accessing immunization services.

### Long waiting times at health facilities

Some caregivers reported arriving very early in the morning at health facilities, having sometimes walked long distances, and then leaving the facility very late in the evening. In some cases, staying for long periods at health facilities raised concerns from spouses and eventually undermined spousal support for immunization. In one community in Mubende district, participants reported that whereas caregivers arrived early at the facility, they had to wait for health workers who sometimes arrived late, which creates build-ups of long queues. Sometimes, health workers were reported to send away caregivers who had waited in the queue because the health worker claimed that they were tired. With such experiences, the caregivers never found it prudent to return to the health facility for services.

> *“Sometimes you can go at 8:00am and return at 5:00pm because the line is very long like from here up to down that side. At times you go back [home] because the health worker has said she is tired; “You will come back another day”*. ***IDI, Caregiver, Mubende district***.

In some communities in Wakiso district, caregivers reported that sometimes the health workers were required to wait until a certain number of caregivers are available to conduct group health education before the immunization. While the health workers acknowledged that waiting too long at the facility sometimes discouraged some caregivers, they argued that arrangements for group health education helped to leverage the fewer health workers available at the facility, who are also required to fill HMIS records and observe the children for any possible adverse events following immunization.

> *“We wait for about 5 to 10 mothers to come before giving them the health education talk. After educating them, I fill in the chart and the register, and after filling those charts and register, I start injecting the child. After injecting the child, I tell the mother to sit there for like 10 to 15 minutes to see whether the child is in good condition. Then, after 15 minutes, I tell her to go home”*. ***KII, Health Worker, Wakiso district***.

In one village in Kasese district, caregivers reported that health workers open the health facility late and close early. This affected the immunization uptake because some mothers could not be attended to given the limited time allocated for immunization.

> *“When you come at 3 pm, you don’t find anyone here. Health workers come from midday up to 2 pm. When it comes to 3 pm and above, there’s no one*.*”*. ***IDI, Caregiver, Kasese district***.

### Poor health worker attitudes

In some communities, caregivers reported that health workers are rude and sometimes they felt embarrassed by the way they were treated by the health workers, which undermined their confidence and self-worth. If a health worker was rude during a particular health visit, the caregivers could not return for vaccination when required, and this contributed to high numbers of under-immunized children.

> *“Some health workers are nice, and others are not nice, they speak rudely. They can abuse us that we villagers and don’t understand*.*”* ***IDI, Caregiver, Mubende district***.

### Costs incurred to access immunization services

In Uganda, immunization services are provided free of charge in government facilities. However, there are indirect and sometimes informal costs that have been reported to constrain uptake of immunization services. The most reported cost was transportation to access distant health facilities. Other costs reported by caregivers included being charged for the immunization cards and health workers demanding money and in-kind goods before providing services or for the child to be prioritized during the immunization session. Caregivers reported that in one facility in Wakiso district, an immunization card for a new mother and replacement after loss ranged between 1,000 [0.27 USD] and 2,000 shillings [0.54 USD] at both public facilities and private health facilities. This practice was also reported in Mubende and Kasese areas as one respondent noted in the quotation below;

> *“I feel so bad when sometimes I come for vaccination, but I do not have money to buy a card for my child. I don’t have any source of income and have no money to buy an immunization card for my child, and even if I had some money, it would be for buying food because I can’t take my child for vaccination when I don’t have food at home*.*”* ***IDI, Caregiver, Kasese district***.
>
> *“But when you are going, you must take something for the health worker, to provide services for the child. Anything like money or something that the health worker will eat. You can offer chicken, or eggs for the child to be immunized. You give her what you have but you must estimate the amount that they can take. You need a minimum of 2,000* [0.54 USD]. *It is the culture; you just get your 2,000 shillings and put it in the middle of the child’s immunization card and hand it over to the health worker because she also does not allow you to hand the money to her when the rest are seeing. So, when you give [it to] her, she will open the card and see the money, remove it when the rest are not seeing*.*”* ***IDI, Caregiver, Mubende***.

The quotations reflect the struggle that caregivers face in determining priorities for the meagre resources they may have, and when faced with a choice problem, taking a child for vaccination becomes secondary. However, in the interactions with the health workers, it was clarified that the charge for the card was typically associated with administrative tasks such as transferring information from existing records or documenting caregiver-reported details on to new child health cards. Although not officially sanctioned, health workers perceive this charge not only as a means to support these documentation processes, but also as a strategy to encourage caregivers to take greater responsibility in maintaining and presenting the child immunization cards during health facility visits.

#### 3. Vaccine stock-outs

Across the high-risk communities, vaccine stockouts were reported as a major reason hindering access to immunization. Caregivers mentioned that sometimes they found that specific vaccines were out of stock and health workers would either refer them to other health facilities, or they would advise them to come after a particular period when they anticipated that vaccines would have be available. In some situations, stockout of a single antigen created a negative demand for other antigens even when they were in stock. For example, when caregivers found BCG out of stock and DPT was available, the BGG stockout created misconceptions among caregivers who assumed every other antigen was unavailable. Consequently, some caregivers chose not to visit the facility, further affecting vaccine uptake. The influence of vaccine availability on uptake was expressed by a respondent in Kasese as follows;

> *“I desire to take my child for immunization, but the most disappointing thing is that you leave your work undone at home, you rush to the facility and when you reach, you find no vaccine. You feel very disappointed and demoralized to take your child another day*.*”* ***IDI, Caregiver, Kasese district***.
>
> *“I have ever taken him[child], and he was immunized the first time. When I went back the second time, I was told that there was no vaccine to immunize him. When I took him for the third time, they said they had not brought the vaccine yet. I went back again for the fourth time; they said that they had not brought the vaccine. On the fifth time, they injected him twice on the thigh, and one on the arm, here, and here [showing the injection sites]”*. ***IDI, Caregiver, Mubende district***.

### Demand-side related barriers

#### 4. Fear of adverse events following immunization

Respondents highlight how post-vaccination effects – such as prolonged crying or discomfort in infants, reported fevers, swelling and pain at the sites of injection, and sometimes lameness, and perceived fears of death – influence household dynamics and caregiver decision-making. In some cases, mothers report being blamed or even asked to leave the room by their spouses when the baby cries persistently overnight following vaccination. These reactions not only cause emotional distress but also limit the mother’s ability to engage in other responsibilities until the child recovers. Sometimes families have discontinued vaccination for successive antigens for fear of adverse effects following their previous experiences. These fears were exacerbated by inadequate information available to the caregivers on the likely adverse events to expect and how to manage them. Sometimes the health workers never provided enough information to the caregiver on what to expect and how to manage those adverse events. These negatively affect decisions to immunize their children. One mother reported that her child became lame because of immunization.

> *“My husband is hesitant about immunization because I took this child, and he got polio, and the leg was bent from behind. He is the only one I took for immunization and after being immunized he became lame, and the father said don’t take him back for immunization. So, I stopped*.*”* ***IDI, Caregiver, Mubende district***.

#### 5. Limited partner involvement and support for immunization

Respondents reported limited spousal support while seeking immunization services. This was exhibited through refusal to give financial support to facilitate transport to the health facility and physical assault when mothers sought immunization services without spousal approval. These sentiments were confirmed by health workers who reported that women expressed such concerns during their interactions at the health facility.

*“I went in the morning and spent the whole day. I realized that it was getting late, and I decided to come back home without immunization because if I came late, my husband would beat me. I have never gone back”*. ***IDI, Caregiver, Mubende district***.

*“*…*you can find a mother who wants to immunize, but a father doesn’t want, and the mother comes and says, Nurse, you give me the injection early, I want to go back when the man has not come back*.*”* ***KII, Health worker, Wakiso district***.

#### 6. Myths, misconceptions, and beliefs

Several myths and misconceptions that discourage caregivers from seeking immunization services were reported. These included cultural beliefs, religious beliefs, community long-held perceptions and beliefs, perceived side effects, vaccine mistrust, and misinformation about immunization. Some religions prohibit their members to participate in programmes that require any form of registration of bio data details because they associate it with some misfortunes and others do not believe in giving medicines to children who are not sick. There are also beliefs about side-effects due to administering expired vaccines.

> *“I was going to take my child, but when I inquired from the women that were coming from immunization, I was told, that there are times when they immunize children, and they are all well, but there are other times when the children get sick. Children get swollen hands, and their bodies get weak*.*”* ***IDI, Caregiver, Mubende district***.
>
> *“There are some parents I contacted, and they said those vaccines they put in their children are microchips. They have the evil number 666 [mentioned to be the mark of the devil in the bible], and the whites are on the other side monitoring them after they insert a microchip in your body. So, I asked them where they got that information from, and they said that they have the Holy Spirit that showed them these things*.*”* ***KII, VHT member, Mubende district***.

#### 7. Home births with or without traditional birth attendants

Despite the ban of traditional birth attendants (TBAs) in Uganda in 2010, the practice continues to be widespread within many communities. Home births including deliveries assisted by traditional birth attendants are reportedly influenced by both cultural and structural factors. Culturally, childbirth is often perceived as a spiritual process that requires more than biomedical expertise – many women believe that TBAs possess the spiritual and experiential knowledge necessary to navigate childbirth and address complications. Structurally, long distances to health facilities and persistent transport challenges further discourage facility-based deliveries. Additionally, some mothers reported negative experiences when seeking postnatal services after delivering at home or with TBAs. There were reported instances of mothers being mishandled or stigmatized by health workers, which may reinforce reluctance to seek vaccination services. This view was reinforced by responses from health workers who reported that children who are delivered outside of the health facility do not often receive immunization services, sometimes fearing reprimand by health workers or to avoid disclosure of places where they delivered from, resulting in children missing the essential birth dose vaccines required.

> *“Some mothers use [TBAs] and some of them deliver by themselves in their homes. They proudly say how well they manage to conduct deliveries at home*.*”* ***KII, Health Worker, Mubende district***.

## Discussion

This was a qualitative rapid assessment of the barriers to utilization of immunization services in three high-burden districts in Uganda. We found that barriers to immunization were both on the demand and supply side. The study findings clearly reflect the four domains of the Behavioral and Social Drivers (BeSD) of vaccination, namely: a) thinking and feeling about vaccines; b) social processes that drive or inhibit vaccination; c) motivation (or hesitancy) to seek vaccination; and d) practical issues involved in seeking and receiving vaccination [11].

Our study found that distance and transport related challenges including poor roads, hilly terrain, and costs constrain access to immunization services. This finding agrees with other studies that find that majority of unvaccinated children live in the inner communities of Sub-Saharan African countries that are difficult to access by healthcare workers on account of poor roads and long distances to immunization posts. Caregivers find it difficult to walk or pay for transport to get to the health facility located far away. Proximity to health facilities influenced utilization of immunization services, with ZD children likely to be in places that are distant from the health facility attributed to the complications that caregivers go through to access immunization services [6, 12–14]. Relatedly, Wasswa et al [5], using the Uganda Demographic and Health Survey (UDHS) data, found that the burden of ZD children was disproportionately higher for rural areas compared to urban areas and argued that this was attributed to the easy access to health facilities in urban areas compared to the rural areas. Outreach immunization services may be a key strategy for serving communities with limited geographical access to health facilities [14].

Most caregivers cited inadequate client-centered immunization services, characterized by negative attitudes of health workers towards clients, indirect costs of accessing immunization, unavailability of health workers at some health facilities, and limited operation time for some facilities. These findings are like what has been reported in other studies. For example, Babirye and colleagues reported that long waiting times mainly in public health facilities result in lost productivity time for caregivers, and this creates a negative perception towards immunization services [12, 15]. The long waiting times are exacerbated by the practice of either waiting for a sizeable number of caregivers to arrive at a facility so as to conduct a mass health education before actual vaccination or waiting for a particular number of children before a vial is opened in order to reduce vaccine wastages [16, 17]. In some situations, it has been noted that due to limited health personnel at health facilities, health workers tend to prioritize curative services over preventive services like immunization. These practices increase the hidden expenses/costs associated with vaccination. Besides, some caregivers perceived the clinic environment and health worker attitudes as barriers to vaccination in this study which is consistent with other studies [18, 19]. The issue of poor attitudes of health workers has been found in other studies as an imbedding factor to immunization. When health workers use unpleasant communication towards caregivers, this undermines repeat vaccination at the same health facility [16, 20–22]. The design, management, and delivery of immunization services should be shaped by and respond to the needs of individuals. A person-centered approach is key in immunization, where it systematically considers the perspectives of individuals, families, and communities, and sees them as active participants and partners in quality and effective service delivery [23].

Vaccine stockouts and supply shortages were a key barrier to completion of immunization schedules for children. Vaccine stockouts lead to missed opportunities for immunization and hence contribute to increased Vaccine Preventable Diseases [8, 14]. Vaccine stockouts are generally attributed to challenges with the supply chain, such as facilities lacking logistics and finances to collect vaccines from the District Vaccine Store [6, 12]. Other causes of vaccine stockouts are attributed to breakdown in cold chain equipment, underestimation of orders at quantification and forecasting, delays in distributing vaccines as per schedule, lack of storage space, and budget constraints [6, 7, 12]. However, UNEPI, with support from implementing partners, has focused on streamlining the vaccine supply chain system, including streamlining mandates and roles for forecasting, quantification, procurement, storage and transportation across the Ministry of Health and National Medical Stores, and there is evidence that these efforts have started paying off [24].

This study found that fear of AEFIs was one the key constraints to uptake of immunization services. The fear for AEFIs generally arises from misinformation (or misleading information), long-held negative perceptions which may be based on cultural, religious, or community beliefs, and inadequate information provided by health workers to the caregivers on expected side effects of vaccination including how the side effects manifest and how they should be managed [14]. Some participants in the current study had experienced vaccine side effects such as fever or excessive crying after vaccination. Although most adverse events related to vaccination are minor, they can lead to either defaulting or delaying subsequent immunizations. The contribution of AEFIs to vaccine hesitancy has been reported in other studies in low- and middle-income countries’ settings [13, 25]. Improved levels of immunization could be achieved if adequate and clear information and education are provided [12]. This has been shown in studies in Gambia and Senegal where women mentioned that they would accept new maternal vaccines if they are sensitized beforehand about any potential risks and benefits to them and their babies [25]. Community leaders and gatekeepers must also be engaged in addressing misinformation and negative perceptions [16].

Partner involvement is very key for decisions on whether or not utilize health care services and a large body of literature on this has been in the area of maternal health and HIV/AIDS [26–28]. Partner involvement and support have also been found to be critical for uptake of childhood immunization services. Partner involvement and support is exemplified by among others; joint decision-making to or not to vaccinate a child, and provision of financial support to cover for transport where necessary, etc. [29–32]. Across the regions, men’s limited engagement and involvement in ensuring their children receive vaccines emerge as a gender specific barrier [33]. Limited male involvement in child health can be understood within the broader socioeconomic context of patriarchal communities. In these settings, women were traditionally viewed as the primary caregivers and are expected to manage health-related matters within the households. Men on the other hand, are often positioned as breadwinners and key decision-makers, with limited engagement in day-to-day caregiving activities [34, 35]. This division of roles influenced the extent to which men participate in vaccination-related decisions and support. In our study, the gender dynamic is further exacerbated by the fact that fathers refuse to provide transport funds to the women to take children for vaccination. A previous study in Uganda found that convictions of the caretakers, self-efficacy, and the supportive or non-supportive role of significant others influenced the involvement or non-involvement of parents in childhood immunization [6, 12]. With regard to decision-making, many cultural settings believe the child belongs to a man who has the ultimate power to decide whether their child should be vaccinated and this is largely influenced by long-held gender stereotypes[32]. Continuous and deliberate provision of information and education to address the gender dynamics coupled with women empowerment programs are apparent to overcome these structural constraints [32].

Consistent with other studies, this study found that myths, misconceptions and beliefs were a crucial challenge that undermined vaccination uptake. The common concerns revolve around the ingredients or composition of the vaccine, propaganda about ‘hidden interests’ of vaccine manufacturers in developed countries; the perceived negative impact of vaccines on fertility; and the religious misinformation about vaccination and the ‘666’ biblical number, etc. [6, 17, 36, 37]. Vaccine ‘infodemic, myths, perceptions, and conspiracies greatly undermine public trust and confidence in vaccines and constrains uptake, hence generating the need for continuous stakeholder engagements guided with adequate information to counter the misinformation [38, 39]. Caregivers should also be provided with information on the benefits of childhood vaccination, as a mechanism to counter misinformation [40].

Finally, home-based deliveries either with or without TBAs undermine uptake of immunization services more especially for vaccines that are expected to be administered immediately after birth. UDHS 2022 reported that 91% of mothers deliver at health facilities, implying that the remaining 9% either deliver at home or at the TBA [2, 3]. Complete vaccination is positively related to maternal health care service utilization including antenatal care and skilled birth attendance, delivery in a public or private health facility compared to being delivered at home [25]. Campaigns to increase institutional or facility-based deliveries are key in reducing non or under-vaccination of children but require addressing the underlying factors that constrain mothers from delivering at health facilities [41]. Given the persistent structural barriers—such as long distances to health facilities, poor road infrastructure, and limited transport options—that drive women to seek delivery services from TBAs, there is value in exploring the potential role of TBAs in facilitating referrals for postnatal services, including vaccination. In many communities, TBAs are trusted figures who hold cultural and spiritual significance in the birthing process. Leveraging their position to promote timely vaccination could help bridge the gap between informal and formal health systems. While this approach may raise concerns about standardization and oversight, it offers a feasible interim strategy in contexts where addressing structural challenges requires long-term investment. Moreover, engaging TBAs in referral pathways could: Improve early postnatal contact with health facilities; enhance community trust in vaccination programs; and reduce missed opportunities for immunization following homebirths. This strategy would need to be carefully designed to ensure TBAs are equipped with accurate information, supported by health systems, and integrated into broader maternal and child health frameworks.

While our study has elicited the key barriers to uptake of immunization for high burden ZD and UI children communities, it may not have captured the full range of insights given the rapid nature of this assessment. Secondly, the results should be interpreted within the context of the study communities and may not necessarily be applied to all ZD /UI children in the general population. Thirdly, data elicited from caregivers may have been subject to recall particularly in situations where immunization cards were not available for reference in which case a caregiver was asked to self-report, and this may have been prone to recall as well as social desirability biases. However, this limitation was minimized by ensuring the interviewers clearly state the rationale and benefits of the study findings to the participants as a way to elicit valid responses. The learning hub has conducted additional rounds of in-depth qualitative inquiries particularly with caregivers of ZD children whose results are expected to provide additional nuances on immunization uptake once shared in subsequent manuscripts.

## Conclusion

Understanding the barriers to immunization uptake among zero-dose and under-immunized children is a critical foundation for strengthening vaccination coverage and reducing the incidence of VPDs in Uganda. This rapid assessment conducted in Kasese, Mubende, and Wakiso districts highlights both demand and supply side challenges that hinder access and utilization of immunization services. Addressing these barriers requires coordinated, multi-sectoral approaches that are both comprehensive and contextually grounded. Tailored strategies that engage communities, enhance service delivery, and prioritize equity are essential to reaching underserved populations, particularly those in high-burden and hard-to-reach areas and ensuring that no child is left behind in Uganda’s immunization efforts.

## Supporting information

Ethical approveal and consent form

## Data Availability

All the data pertaining to this article is available, on request at the Infectious Disease Research Institute, Kampala Head Office Plot 2C Nakasero Hill Road, P.O. Box 7475 Kampala, Uganda Phone: (+256) 312 281-479.

## Acknowledgments

The authors extend their sincere gratitude to the entire team at Uganda National Expanded Programme for Immunization (UNEPI), Ministry of Health, as well as teams from UNICEF, CHAI, WHO, and other partners for the support during the implementation of the study. We also extend our appreciation to the Local Government authorities of Kasese, Mubende, and Wakiso districts particularly the District Health Team (DHT and VHT members for supporting the implementation of the study. The authors also appreciate the study participants for supporting the implementation of the study.

This study was conducted under the auspices of the Uganda Zero-dose Learning Hub, a collaboration between the Infectious Diseases Research Collaboration (IDRC), PATH Uganda, and Makerere University School of Public Health (MakSPH), with funding from the Global Alliance for Vaccines Initiative (Gavi) (Uganda Learning Hub, MEL:12564223). The funders had no role in the design, implementation of the study and the decision to publish the findings. The views and opinions expressed in this study reflect the findings from the study and the views of the authors and not views of the funding organization.

## Data availability statement

The data for this study are available from the corresponding author upon reasonable request.

## Authors’ contributions

**Study conceptualization and design:** Emmanuel Mugisha, PhD (EM), Shadiah Nanteza Mugizi, MPH (SNM), Deogratias Agaba, MPH (DA) and Jacqueline Anena, MPH (JA) and Joaniter I. Nankabirwa, PhD (JIN).

**Study implementation and data collection support:** EM, SNM, JIN, Fred Isaasi, MSC (FI), David Wafula, MPH (DW), Betty Mirembe, MD (BM), and Susan Nayiga, PhD (SN).

**Data transcription, cleaning and management:** FI, DW, SNM, and Miriam Kayendeke, MSC (MK).

**Data analysis:** EM, SNM), JIN, SN, AK, BM, DA, JA, Allen Kabagenyi (AK), Peter Waiswa (PW); Moses R. Kamya, PhD (MRK); Chrispus Mayora, PhD (CM), Patrick Albert Ipola, MPH (PIA), Paul Katamba, MPH (PK), Carol Kamya, MPH (CK). Faith Namugaya, MPH (FN), and MK

**Writing initial manuscript draft:** EM, SNM, CM, and BM.

**Review and revision of manuscript:** EM, CM, SNM, JIN, SN, AK, BM, DA, JA, AK, PW, CK, FM, MRK, PK, PIA, FI, DW, FN, and MK.

All authors read and approved the final manuscript and met the ICMJE criteria for authorship.

