## Supplementary material for "Barriers to reaching the Zero-Dose and Under Immunized Children in Uganda: a qualitative rapid assessment": Ethical approveal and consent form

Uganda National Council for Science and Technology
*(Established by Act of Parliament of the Republic of Uganda)*
UNCST

**Our Ref: HS3011ES** **22 September 2023**

Kamya Moses
Infectious Disease Research Collaboration (IDRC) , Uganda
**Kampala**

**Re: Research Approval: Optimising strategies to reach zero-dose, under-immunized children, and missed communities in Uganda.**

I am pleased to inform you that on **22/09/2023**, the Uganda National Council for Science and Technology (UNCST) approved the above referenced research project. The Approval of the research project is for the period of **22/09/2023** to **22/09/2026**.

Your research registration number with the UNCST is **HS3011ES**. Please, cite this number in all your future correspondences with UNCST in respect of the above research project. As the Principal Investigator of the research project, you are responsible for fulfilling the following requirements of approval:

1. Keeping all co-investigators informed of the status of the research.

2. Submitting all changes, amendments, and addenda to the research protocol or the consent form (where applicable) to the designated Research Ethics Committee (REC) or Lead Agency for re-review and approval **prior** to the activation of the changes. UNCST must be notified of the approved changes within five working days.

3. For clinical trials, all serious adverse events must be reported promptly to the designated local REC for review with copies to the National Drug Authority and a notification to the UNCST.

4. Unanticipated problems involving risks to research participants or other must be reported promptly to the UNCST. New information that becomes available which could change the risk/benefit ratio must be submitted promptly for UNCST notification after review by the REC.

5. Only approved study procedures are to be implemented. The UNCST may conduct impromptu audits of all study records.

6. An annual progress report and approval letter of continuation from the REC must be submitted electronically to UNCST. Failure to do so may result in termination of the research project.

Please note that this approval includes all study related tools submitted as part of the application as shown below:

| No. | Document Title | Language | Version Number | Version Date |
|---|---|---|---|---|
| 1 | Risk Management Plan | English | 1.0 | 11 May 2023 |
| 2 | Appendix B_ KII topic guide VHTs and Community Leaders | English | 2.0 | 16 June 2023 |
| 3 | Appendix C_KII Topic guide_National and sub national level | English | 2.0 | 16 June 2023 |
| 4 | ppendix D_KII Topic guide_Service Providers | English | 2.0 | 16 June 2023 |
| 5 | Appendix F_IDI Topic guide Mother Primary care providers | English | 2.0 | 16 June 2023 |
| 6 | Appendix G_TOOL Target Survey questionnaire | English | 2.0 | 16 June 2023 |
| 7 | HFA-Assessment tool | English | 2.0 | 16 June 2023 |
| 8 | Community Engagement Plan | English | 1.0 | 11 May 2023 |
| 9 | Appendix A GAVI KII IC_VHTs and Community leaders | English | 2.0 | 16 June 2023 |
| 10 | Appendix E GAVI IDI IC | English | 2.0 | 16 June 2023 |
| 11 | Appendix H GAVI community survey IC | English | 2.0 | 16 June 2023 |
| 12 | Appendix M GAVI KII IC Sub national and national stakeholders | English | 2.0 | 16 June 2023 |
| 13 | Appendix J GAVI HFA IC | English | 2.0 | 16 June 2023 |
| 14 | Appendix E GAVI IDI IC | Luganda | 2.0 | 16 June 2023 |
| 15 | Appendix M GAVI KII IC Sub national and national stakeholders | Luganda | 2.0 | 16 June 2023 |
| 16 | Appendix H GAVI community survey IC_V2.0 16 June 2023 | Luganda | 2.0 | 16 June 2023 |
| 17 | Appendix A GAVI KII IC_VHTs and Community leaders | Luganda | 2.0 | 16 June 2023 |
| 18 | Appendix A GAVI KII IC_VHTs and Community leaders | Lhukozo | 2.0 | 16 June 2023 |
| 19 | Appendix E GAVI IDI IC | Lhukozo | 2.0 | 16 June 2023 |
| 20 | Appendix H GAVI community survey IC | Lhukozo | 2.0 | 16 June 2023 |
| 21 | Appendix M GAVI KII IC Sub national and national stakeholders | Lhukozo | 2.0 | 16 June 2023 |
| 22 | Appendix A GAVI KII IC_VHTs and Community leaders | Rutooro | 2.0 | 16 June 2023 |
| 23 | Appendix E GAVI IDI IC | Rutooro | 2.0 | 16 June 2023 |
| 24 | Appendix H GAVI community survey IC | Rutooro | 2.0 | 16 June 2023 |
| 25 | Appendix M GAVI KII IC Sub national and national stakeholders | Rutooro | 2.0 | 16 June 2023 |
| 26 | Project Proposal | English | 2.0 | |
| 27 | Approval Letter | English | | |
| 28 | Administrative Clearance | English | | |

Yours sincerely,

Hellen Opolot

*LOCATION/CORRESPONDENCE*

*Plot 6 Kimera Road, Ntinda*
*P.O. Box 6884*
*KAMPALA, UGANDA*

*COMMUNICATION*

**WEBSITE: http://www.uncst.go.ug**

### MAKEREREUNIVERSITY

P.O. Box 7072
Kampala UGANDA


Website: www.sph.mak.ac.ug


#### COLLEGE OF HEALTH SCIENCES
#### SCHOOL OF PUBLIC HEALTH

**Research and Ethics Committee**

**01/08/2023**

To: Kamya Moses

**Type:** Initial Review

**Re: SPH-2023-428: Optimizing strategies to reach zero-dose, under-immunized children, and missed communities in Uganda**

I am pleased to inform you that at the **Fast track meeting** convened meeting on **23/05/2023**, the MAK School of Public Health REC (SPHREC) meeting voted to approve the above referenced application.
Approval of the research is for the period of **01/08/2023** to **01/08/2024.**

As Principal Investigator of the research, you are responsible for fulfilling the following requirements of approval:

1. All co-investigators must be kept informed of the status of the research.
2. Changes, amendments, and addenda to the protocol or the consent form must be submitted to the REC for re-review and approval **prior** to the activation of the changes.
3. Reports of unanticipated problems involving risks to participants or any new information which could change the risk benefit: ratio must be submitted to the REC.
4. Only approved consent forms are to be used in the enrollment of participants. All consent forms signed by participants and/or witnesses should be retained on file. The REC may conduct audits of all study records, and consent documentation may be part of such audits.
5. Continuing review application must be submitted to the REC **eight weeks** prior to the expiration date of **01/08/2024** in order to continue the study beyond the approved period. Failure to submit a continuing review application in a timely fashion may result in suspension or termination of the study.
6. The REC application number assigned to the research should be cited in any correspondence with the REC of record.
7. You are required to register the research protocol with the Uganda National Council for Science and Technology (UNCST) for final clearance to undertake the study in Uganda.

The following is the list of all documents approved in this application by MAK School of Public Health REC (SPHREC):

| No. | Document Title | Language | Version Number | Version Date |
|---|---|---|---|---|
| 1 | 11_Appendix M GAVI KII IC Sub national and national stakeholders _V2.0 16 June 2023 | Rutooro | 2.0 | 2023-06-16 |
| 2 | 9b_Appendix H GAVI community survey IC_V2.0 16 June 2023 | Rutooro | 2.0 | 2023-06-16 |
| 3 | 6b_Appendix E GAVI IDI IC_V2.0 16 June 2023 | Rutooro | 2.0 | 2023-06-16 |
| 4 | 2b_Appendix A GAVI KII IC_VHTs and Community leaders_16 June 2023 | Rutooro | 2.0 | 2023-06-16 |
| 5 | 11_Appendix M GAVI KII IC Sub national and national stakeholders _V2.0 16 June 2023 | Luganda | 2.0 | 2023-06-16 |
| 6 | 9b_Appendix H GAVI community survey IC_V2.0 16 June 2023 | Luganda | 2.0 | 2023-06-16 |
| 7 | 6b_Appendix E GAVI IDI IC_V2.0 16 June 2023 | Luganda | 2.0 | 2023-06-16 |
| 8 | 2b_Appendix A GAVI KII IC_VHTs and Community leaders_16 June 2023 | Luganda | 2.0 | 2023-06-16 |
| 9 | 11_Appendix M GAVI KII IC Sub national and national stakeholders _V2.0 16 June 2023 | Lhukozo | 2.0 | 2023-06-16 |
| 10 | 9b_Appendix H GAVI community survey IC_V2.0 16 June 2023 | Lhukozo | 2.0 | 2023-06-16 |
| 11 | 6b_Appendix E GAVI IDI IC_V2.0 16 June 2023_ | Lhukozo | 2.0 | 2023-06-16 |
| 12 | 2b_Appendix A GAVI KII IC_VHTs and Community leaders_16 June 2023 | Lhukozo | 2.0 | 2023-06-16 |
| 13 | 9b_Appendix H GAVI community survey IC_V2.0 16 June 2023_clean | English | 2.0 | 2023-06-16 |
| 14 | Risk Mitigation Plan | English | 1.0 | 2023-05-11 |
| 15 | Community Engagement Plan | English | 1.0 | 2023-05-11 |
| 16 | 11_Appendix M GAVI KII IC Sub national and national stakeholders _V2.0 16 June 2023 | English | 2.0 | 2023-06-16 |
| 17 | 12_HFA-Assessment tool V2.0 dated 16 June 2023 | English | 2.0 | 2023-06-16 |
| 18 | 5_Appendix D_KII Topic guide_Service Providers_V2.0 16 June 2023 | English | 2.0 | 2023-06-16 |
| 19 | 4_Appendix C_KII Topic guide_National and sub national level_V2.0 16 June 2023 | English | 2.0 | 2023-06-16 |
| 20 | 3_Appendix B_ KII topic guide VHTs and | English | 2.0 | 2023-06-16 |

| | | | | |
|---|---|---|---|---|
| | Community Leaders_V2.0 16 June 2023 | | | |
| 21 | 8 Appendix G_TOOL_Target Survey questionaire_V2.0 16 June 2023 | English | 2.0 | 2023-06-16 |
| 22 | Data collection tools | English | 2.0 | 2023-- |
| 23 | 10b_Appendix J GAVI HFA IC_V2.0 dated 16 June 2023_clean | English | 2.0 | 2023-06-15 |
| 24 | 10a_Appendix J GAVI HFA IC_V2.0 dated 16 June 2023_track changes | English | 2.0 | 2023-06-16 |
| 25 | 9a_Appendix H GAVI community survey IC_V2.0 16 June 2023_Track changes | English | 2.0 | 2023-06-16 |
| 26 | 6b_Appendix E GAVI IDI IC_V2.0 16 June 2023_clean | English | 2.0 | 2023-06-16 |
| 27 | 6a_Appendix E GAVI IDI IC_V2.0 16 June 2023_Track changes | English | 2.0 | 2023-06-16 |
| 28 | 2b_Appendix A GAVI KII IC_VHTs and Community leaders_16 June 2023_clean | English | 2.0 | 2023-06-16 |
| 29 | 2a_Appendix A GAVI KII IC_VHTs and Community leaders_16 June 2023_Track changes | English | 2.0 | 2023-06-16 |
| 30 | 1b_Gavi LH research protocol_clean- ver 2.0 16-06-23 | English | 2.0 | 2023-06-16 |
| 31 | Data sharing plan | English | 1.0 | 2023-05-02 |
| 32 | Joaniter Nankabirwa- CV | English | 1.0 | 2023-04-20 |
| 33 | Nayiga Suzan | English | 1.0 | 2023-04-20 |
| 34 | Waisswa Peter | English | 1.0 | 2023-04-20 |
| 35 | Carol Kamya CV | English | 1.0 | 2023-04-20 |
| 36 | Prof Kamya Moses-CV | English | 1.0 | 2022-09-20 |

Yours Sincerely

Joseph Kagaayi
For: MAK School of Public Health REC (SPHREC)

MAKERERE UNIVERSITY
SCHOOL OF PUBLIC HEALTH
APPROVED
VALID UNTIL
★ 01 AUG 2024 ★
RESEARCH & ETHICS COMMITTEE
P.O. BOX 7072, KAMPALA

#### APPENDIX A
#### Key Informant Interviews Participation Informed Consent Form
#### (Village Health Team members and Community leaders)

| | |
|---|---|
| **Protocol Title:** | Optimizing strategies to reach zero-dose, under-immunized children, and missed communities in Uganda. |
| **Funding Source:** | Gavi, The Vaccine Alliance |
| **MakSPH-REC Number:** | SPH-2023-428 |
| **UNCST Number:** | HS3011ES |

**Ugandan Principal Investigator**: Prof. Moses R Kamya, MBChB, MMed, PhD

**Version Date:** 2.0 16th June 2023

##### INTRODUCTION
My name is ……………………………….., I am a research assistant working on the research study entitled "Optimizing strategies to reach zero-dose, under-immunized children, and missed communities in Uganda". You are being asked to participate in this research because you are key stakeholder for immunization services in Uganda. This study is being done by researchers from Infectious Diseases Research Collaboration (IDRC), Makerere University (MU), and PATH Uganda, and is sponsored by Gavi, The Vaccine Alliance. Before you decide if you want to participate in this study, we want you to know as much as possible about the study.

This consent form gives you information about what is involved in participation in this research study. The information in this form will help you decide if you want to participate in the study. You can ask questions about this study at any time. If you agree to participate in this study, we will ask you to sign the consent form. You will get a copy of this form to keep.

##### WHY IS THIS STUDY BEING DONE?
The information collected as part of the key informant interviews will help us get practical and locally relevant strategies for increasing access to immunization services for zero doze and under immunized children in Uganda. The interviews will also provide insights on who the ZDC, UI children, and missed communities are and the reasons for this, the different strategies and interventions that have been implemented to improve vaccination for the ZDC, UI children, and missed communities as well as areas for improvement.

##### WHAT WILL HAPPEN IF I AGREE TO PARTICIPATE IN THE STUDY?
If you agree to participate in this study, the following will happen:

a. Our study team will talk to you about the study and ask you some questions to determine if you are interested and eligible to join the study.
b. If you are interested and eligible to join the study, we will ask that you sign this consent form.

MAKERERE UNIVERSITY
SCHOOL OF PUBLIC HEALTH
APPROVED
VALID UNTIL
01 AUG 2024
RESEARCH & ETHICS COMMITTEE
P O BOX 7072, KAMPALA

c. After the consent form is signed, the study team member will ask you some brief questions about immunization services in immunization services in Uganda. Questions will be centered around:
   - The existing gaps in immunization
   - Experiences in delivering/supporting immunization services
   - What has worked well and the challenges to immunization in Uganda.
   - Strategies and interventions that have been undertaken to increase immunization coverage among the zero dose children, under immunized children, and missed communities.
   - Areas for improvement in immunization services.

We will take notes of the discussion and will record the interview using a digital voice recorder. Afterwards, we will write the ideas from the audio recordings in a computer to allow for analysis.

**HOW LONG WILL I BE IN THIS STUDY?**

Overall, the study will last for about three years. Today, the interview will last about 30-90 minutes.

**CAN I STOP BEING IN THE STUDY?**

Yes. You can decide to stop being in the study at any time. Tell the research team member if you are thinking about stopping or deciding to stop.

**WHAT RISKS CAN I EXPECT FROM BEING IN THE STUDY?**

Topic guides: Answering some questions may make you feel uncomfortable. You may refuse to answer any question or stop at any time. This will not result in any penalty or other disadvantage for you.

Confidentiality: Other people may learn that you are part of this study because we are talking to you. However, we will not allow people who are not working for the study to see any study information we collect from you.

**ARE THERE BENEFITS TO TAKING PART IN THIS STUDY?**

There will be no direct benefits to you, however, the information we get from this study might help Uganda and other countries to decide how to reach zero-dose and under-immunized children.

**WHAT OTHER CHOICES DO I HAVE IF I DO NOT WANT TO TAKE PART IN THIS STUDY?**

You are free to decide whether you want to be in this study. If you decide you do not want to be in this study or decide to stop being in the study at any time and for any reason, this will not affect you.

**WILL MY INFORMATION BE KEPT PRIVATE?**

Other people may learn that you are part of this study because we are talking to you; however, we will not allow people who are not working for the study to see any information we are collecting from you. The universities and research organizations running this study are not allowed to let others know the identity of the people in the study. The records for the study will be kept in a locked office and will only be seen by study workers. Your name will not be written in any reports based on this research. People or organizations which may review your records include: the Uganda Ministry of Health, Makerere University School of Public Health research ethics committee and study staff.

MAKERERE UNIVERSITY
SCHOOL OF PUBLIC HEALTH
APPROVED
VALID UNTIL
★ 01 AUG 2024 ★
RESEARCH/ETHICS COMMITTEE
P O. BOX 7072, KAMPALA

**WHAT ARE THE COSTS TO ME?**
There will be no cost for you to take part in this study.

**WILL I BE PAID FOR TAKING PART IN THIS STUDY?**
You will not be paid for participation in the study. We will be giving you 20,000/= as compensation for the time you spend on study-related activities.

**WHAT ARE MY RIGHTS FOR TAKING PART IN THIS STUDY?**
You are free to decide whether you want to be in this study. You have the right to stop your participation in the study at any time without penalty or loss of benefits to which you are otherwise entitled.

**WHO CAN ANSWER MY QUESTIONS ABOUT THE STUDY?**
Dr. Moses Kamya and any other study staff can answer questions you have about the study. You may call Dr Moses on mobile: 031-2-281479/ 0752 900 012. We will give you free access to a telephone line. You may also contact Dr Kagaayi Joseph (0702 444154) at Makerere University School of Public Health Higher Degrees Research and Ethics Committee which approved this study for questions about participants' rights and research-related harm.

**CONSENT: WHAT YOUR SIGNATURE OR THUMBPRINT MEANS**
PARTICIPATION IN RESEARCH IS VOLUNTARY. You have the right to decline to participate or to withdraw from the study at any point in this study without penalty or loss of benefits to which you are otherwise entitled. A copy of this consent form will be given to you. Your signature or thumbprint below means that you have had this study explained to you. Your signature or thumbprint below means you have had the opportunity to ask questions and to get answers. If you wish to participate in this study, you should sign or place your thumbprint below.

____________________________________________
Name of Participant (printed)

____________________________________________
Signature/Thumbprint Date

____________________________________________
Name of Study Staff Administering Consent (printed) Position/Title

____________________________________________
Signature of Study Staff Administering Consent Date

*If the participant is unable to read and/or write, an impartial witness must be present during the informed consent discussion. After the written informed consent form is read and explained to the participants. After oral consent to participate in the study and the participants has provided her/his fingerprint, the witness should sign and personally date the consent form. By signing the consent form, the witness attests that the information in the consent form and any other written information was accurately explained to, and apparently understood by, the parent or guardian, and that informed consent was freely given by the participant and parent or guardian.

________________________________________________
Name of Person Witnessing Consent (printed)

________________________________________________
Signature of Person Witnessing Consent                Date

MAKERERE UNIVERSITY
SCHOOL OF PUBLIC HEALTH
APPROVED
VALID UNTIL
★ 01 AUG 2024 ★
RESEARCH & ETHICS COMMITTEE
P. O. BOX 7072, KAMPALA

**APPENDIX E**

**IN-DEPTH INTERVIEWS (IDIs) Participation Informed Consent Form**

| | |
|---|---|
| **Protocol Title:** | Optimizing strategies to reach zero-dose, under-immunized children, and missed communities in Uganda. |
| **Funding Source:** | Gavi, The Vaccine Alliance |
| **MakSPH-REC Number:** | SPH-2023-428 |
| **UNCST Number:** | HS3011ES |

MAKERERE UNIVERSITY SCHOOL OF PUBLIC HEALTH
APPROVED
VALID UNTIL
★ 01 AUG 2024 ★
RESEARCH & ETHICS COMMITTEE
P.O. BOX 7072, KAMPALA

**Ugandan Principal Investigator**: Prof. Moses R Kamya, MBChB, MMed, PhD

**Version Date:** 2.0 dated 16th June 2023

#### INTRODUCTION

My name is ......................................., I am a research assistant working on the research study entitled "Optimizing strategies to reach zero-dose, under-immunized children, and missed communities in Uganda". You are being asked to participate in this research study because you are a key stakeholder for immunization services in Uganda. This study is being done by researchers from Infectious Diseases Research Collaboration (IDRC), Makerere University (MU), and PATH Uganda, and is sponsored by Gavi, The Vaccine Alliance. Before you decide if you want to participate in this study, we want you to know as much as possible about the study.

This consent form gives you information about what is involved in the participation in this research study. The information in this form will help you to decided you want to participation in the study. You can ask questions about this study at any time. If you agree to participate in this study, we will ask you to sign the consent form. You will get a copy of this form to keep.

#### WHY IS THIS STUDY BEING DONE?

The purpose of the in-depth interviews is to help us understand the perspectives of mothers, fathers, and caregivers of zero-dose and under immunized children, and households that are part of communities with high numbers of these children. This information will help us get practical and locally relevant strategies for increasing access to immunization services in these communities.

#### WHAT WILL HAPPEN IF I AGREE TO PARTICIPATE IN THE STUDY?

If you agree to participate in this study, we will ask you questions about your experience receiving immunization services including:

- What has worked well for you, what you liked, and what you did not like when accessing immunization services.
- and areas for improvement?
- The challenges you have faced in accessing services.
- What can be done improve access to immunization services.
- Your experience with interventions designed to increase uptake of immunization among zero-dose and under immunized children.

MAKERERE UNIVERSITY
SCHOOL OF PUBLIC HEALTH
APPROVED
VALID UNTIL
01 AUG 2024
RESEARCH & ETHICS COMMITTEE
P O BOX 7072, KAMPALA

We will take notes of the discussion and will record the interview using a digital voice recorder. Afterwards, we will write the ideas captured in the audio recordings in a computer to allow for analysis.

**HOW LONG WILL I BE IN THIS STUDY?**
Overall, the study will last for about three years. Today, the interview will last about 30-90 minutes.

**CAN I STOP BEING IN THE STUDY?**
Yes. You can decide to stop being in the study at any time. Tell the research team member if you decide to stop.

**WHAT RISKS CAN I EXPECT FROM BEING IN THE STUDY?**
Questionnaires: Answering some questions may make you feel uncomfortable. You may refuse to answer any question or stop at any time. This will not result in any penalty or other disadvantage for you.

Confidentiality: Other people may learn that you are part of this study because we are talking to you. However, we will not allow people who are not working for the study to see any study information we collect from you.

**ARE THERE BENEFITS TO TAKING PART IN THIS STUDY?**
There will be no direct benefits to you, however, the information we get from this study might help Uganda and other countries to decide how to reach and improve immunization services for zero-dose and under-immunized children.

**WHAT OTHER CHOICES DO I HAVE IF I DO NOT WANT TO TAKE PART IN THIS STUDY?**
You are free to decide whether you want to be in this study. If you decide you do not want to be in this study or decide to stop being in the study at any time and for any reason, this will not affect you.

**WILL MY INFORMATION BE KEPT PRIVATE?**
Other people may learn that you are part of this study because we are talking to you; however, we will not allow people who are not working for the study to see any information we are collecting from you. The universities and research organizations running this study are not allowed to let others know the identity of the people in the study. The records for the study will be kept in a locked office and will only be seen by study workers. Your name will not be written in any reports based on this research. People or organizations which may review your records include: the Uganda Ministry of Health, Makerere University School of Public Health research ethics committee and study staff.

**WHAT ARE THE COSTS TO ME?**
There will be no cost for you to take part in this study.

**WILL I BE PAID FOR TAKING PART IN THIS STUDY?**
You will not be paid for participation in the study. We will be giving you 20,000/= as compensation for the time you spend on study-related activities.

#### WHAT ARE MY RIGHTS FOR TAKING PART IN THIS STUDY?

You are free to decide whether you want to be in this study. You have the right to stop your participation in the study at any time without penalty or loss of benefits to which you are otherwise entitled.

#### WHO CAN ANSWER MY QUESTIONS ABOUT THE STUDY?

Dr Moses Kamya and the study staff can answer questions you have about the study. You may call Dr Moses on mobile: 031-2-281479/ 0752 900 012. We will give you free access to a telephone line. You may also contact Dr Kagaayi Joseph (0702 444154) at Makerere University School of Public Health Higher Degrees Research and Ethics Committee which approved this study for questions about participants' rights and research-related harm.

#### CONSENT: WHAT YOUR SIGNATURE OR THUMBPRINT MEANS

PARTICIPATION IN RESEARCH IS VOLUNTARY. You have the right to decline to participate or to withdraw from the study at any point in this study without penalty or loss of benefits to which you are otherwise entitled. A copy of this consent form will be given to you. Your signature or thumbprint below means that you have had this study explained to you. Your signature or thumbprint below means you have had the opportunity to ask questions and to get answers. If you wish to participate in this study, you should sign or place your thumbprint below.

________________________________________

Name of Participant (printed)

________________________________________

Signature/Thumbprint Date

________________________________________

Name of Study Staff Administering Consent (printed) Position/Title

________________________________________

Signature of Study Staff Administering Consent Date

*If the participant is unable to read and/or write, an impartial witness must be present during the informed consent discussion. After the written informed consent form is read and explained to the participants. After oral consent to participate in the study and the participants has provided her/his fingerprint, the witness should sign and personally date the consent form. By signing the consent form, the witness attests that the information in the consent form and any other written information was accurately explained to, and apparently understood by, the parent or guardian, and that informed consent was freely given by the participant and parent or guardian.

________________________________________

Name of Person Witnessing Consent (printed)

________________________________________

Signature of Person Witnessing Consent Date

MAKERERE UNIVERSITY
SCHOOL OF PUBLIC HEALTH
APPROVED
VALID UNTIL
01 AUG 2024
RESEARCH & ETHICS COMMITTEE
P O BOX 7072, KAMPALA

**APPENDIX M**
**Key Informant Interviews Participation Informed Consent Form**
**Sub national and national stakeholders**

| | |
|---|---|
| **Protocol Title:** | Optimizing strategies to reach zero-dose, under-immunized children, and missed communities in Uganda. |
| **Funding Source:** | Gavi, The Vaccine Alliance |
| **MakSPH-REC Number:** | SPH-2023-428 |
| **UNCST Number:** | HS3011ES |

MAKERERE UNIVERSITY SCHOOL OF PUBLIC HEALTH APPROVED VALID UNTIL ★ 01 AUG 2024 ★ RESEARCH & ETHICS COMMITTEE P O BOX 7072, KAMPALA

**Ugandan Principal Investigator:** Prof. Moses R Kamya, MBChB, MMed, PhD

**Version Date:** 2.0 dated 16th June 2023

#### INTRODUCTION

My name is ……………………………………., I am a research assistant working on the research study entitled "Optimizing strategies to reach zero-dose, under-immunized children, and missed communities in Uganda". You are being asked to participate in this research study because you are key stakeholder for immunization services in Uganda. This study is being done by researchers from Infectious Diseases Research Collaboration (IDRC), Makerere University (MU), and PATH Uganda, and is sponsored by Gavi, The Vaccine Alliance. Before you decide if you want to participate in this study, we want you to know as much as possible about the study.

This consent form gives you information about what is involved in participation in this research study. The information in this form will help you decide if you want to participate in the study. You can ask questions about this study at any time. If you agree to participate in this study, we will ask you to sign the consent form. You will get a copy of this form to keep.

#### WHY IS THIS STUDY BEING DONE?

The information collected as part of the key informant interviews will help us get practical and locally relevant strategies for increasing access to immunization services for zero doze and under immunized children in Uganda. The interviews will also provide insights on the different strategies and interventions that have been implemented to improve vaccination for the ZDC, UI children, and missed communities as well as areas for improvement.

#### WHAT WILL HAPPEN IF I AGREE TO PARTICIPATE IN THE STUDY?

If you agree to participate in this study, the following will happen:

a. Our study team will talk to you about the study and ask you some questions to determine if you are interested and eligible to join the study.
b. If you are interested and eligible to join the study, we will ask that you sign this consent form.

c. After the consent form is signed, the study team member will ask you some brief questions about immunization services in immunization services in Uganda. Questions will be centered around:
   - The existing gaps in immunization
   - Experiences in delivering/supporting immunization services
   - What has worked well and the challenges to immunization in Uganda.
   - Strategies and interventions that have been undertaken to increase immunization coverage among the zero dose children, under immunized children, and missed communities.
   - Areas for improvement in immunization services.
   - Immunization data capture systems including the available systems, data captured, data use, tools, and sustainability of systems.

We will take notes of the discussion and will record the interview using a digital voice recorder. Afterwards, we will write the ideas from the audio recordings in a computer to allow for analysis.

**HOW LONG WILL I BE IN THIS STUDY?**
Overall, the study will last for about three years. Today, the interview will last about 30-90 minutes.

**CAN I STOP BEING IN THE STUDY?**
Yes. You can decide to stop being in the study at any time. Tell the research team member if you are thinking about stopping or deciding to stop.

**WHAT RISKS CAN I EXPECT FROM BEING IN THE STUDY?**
Topic guides: Answering some questions may make you feel uncomfortable. You may refuse to answer any question or stop at any time. This will not result in any penalty or other disadvantage for you.

Confidentiality: Other people may learn that you are part of this study because we are talking to you. However, we will not allow people who are not working for the study to see any study information we collect from you.

**ARE THERE BENEFITS TO TAKING PART IN THIS STUDY?**
There will be no direct benefits to you, however, the information we get from this study might help Uganda and other countries to decide how to reach zero-dose and under-immunized children.

**WHAT OTHER CHOICES DO I HAVE IF I DO NOT WANT TO TAKE PART IN THIS STUDY?**
You are free to decide whether you want to be in this study. If you decide you do not want to be in this study or decide to stop being in the study at any time and for any reason, this will not affect you.

MAKERERE UNIVERSITY
SCHOOL OF PUBLIC HEALTH
APPROVED
VALID UNTIL
★ 01 AUG 2024 ★
RESEARCH & ETHICS COMMITTEE
P O BOX 7072, KAMPALA

**WILL MY INFORMATION BE KEPT PRIVATE?**
Other people may learn that you are part of this study because we are talking to you; however, we will not allow people who are not working for the study to see any information we are collecting from you. The universities and research organizations running this study are not allowed to let others know the identity of the people in the study. The records for the study will be kept in a locked office and will only be seen by study workers. Your name will not be written in any reports based on this research. People or organizations which may review your records include: the Uganda Ministry of Health, Makerere University School of Public Health research ethics committee and study staff.

**WHAT ARE THE COSTS TO ME?**
There will be no cost for you to take part in this study.

**WILL I BE PAID FOR TAKING PART IN THIS STUDY?**
You will not be paid for participation in the study. We will be giving you 20.000/= as compensation for the time you spend on study-related activities.

**WHAT ARE MY RIGHTS FOR TAKING PART IN THIS STUDY?**
You are free to decide whether you want to be in this study. You have the right to stop your participation in the study at any time without penalty or loss of benefits to which you are otherwise entitled.

**WHO CAN ANSWER MY QUESTIONS ABOUT THE STUDY?**
Dr Moses Kamya and the study staff can answer questions you have about the study. You may call Dr Moses on mobile: 031-2-281479/ 0752 900 012. We will give you free access to a telephone line. You may also contact Dr Kagaayi Joseph (0702 444154) at Makerere University School of Public Health Higher Degrees Research and Ethics Committee which approved this study for questions about participants' rights and research-related harm.

MAKERERE UNIVERSITY
SCHOOL OF PUBLIC HEALTH
APPROVED
VALID UNTIL
★ 01 AUG 2024 ★
RESEARCH & ETHICS COMMITTEE
P.O. BOX 7072, KAMPALA

**CONSENT: WHAT YOUR SIGNATURE OR THUMBPRINT MEANS**

PARTICIPATION IN RESEARCH IS VOLUNTARY. You have the right to decline to participate or to withdraw from the study at any point in this study without penalty or loss of benefits to which you are otherwise entitled. A copy of this consent form will be given to you. Your signature or thumbprint below means that you have had this study explained to you. Your signature or thumbprint below means you have had the opportunity to ask questions and to get answers. If you wish to participate in this study, you should sign or place your thumbprint below.

________________________________________

Name of Participant (printed)

________________________________________

Signature/Thumbprint Date

________________________________________

Name of Study Staff Administering Consent (printed) Position/Title

________________________________________

Signature of Study Staff Administering Consent Date

*If the participant is unable to read and/or write, an impartial witness must be present during the informed consent discussion. After the written informed consent form is read and explained to the participants. After oral consent to participate in the study and the participants has provided her/his fingerprint, the witness should sign and personally date the consent form. By signing the consent form, the witness attests that the information in the consent form and any other written information was accurately explained to, and apparently understood by, the parent or guardian, and that informed consent was freely given by the participant and parent or guardian.

________________________________________

Name of Person Witnessing Consent (printed)

________________________________________

Signature of Person Witnessing Consent Date

MAKERERE UNIVERSITY
SCHOOL OF PUBLIC HEALTH
APPROVED
VALID UNTIL
★ 01 AUG 2024 ★
RESEARCH & ETHICS COMMITTEE
P O. BOX 7072, KAMPALA

### APPENDIX F: IN DEPTH INTERVIEW TOPIC GUIDE

#### Mothers/Primary care providers

#### PART 1: DEMOGRAPHIC INFORMATION

| | | | |
|---|---|---|---|
| **1. Age** | Years [___\|___] | **5. Highest level of education or qualification achieved**<br>0 = None<br>1 = Primary (P1 — P7)<br>2 = Secondary (S1 — S6)<br>3 = Certificate<br>77 = Other______________ | 4 = Diploma<br>5 = Bachelor's degree<br>88 = Don't know<br>99 = Refused to answer<br>[___\|___] |
| **2. Gender** | 1 = Male<br>2 = Female [___] | | |
| **3. Number of children under your care** | [___\|___] | | |
| **4. Age of youngest child** | Years [___\|___]<br>Months [___\|___] | **6. Primary occupation** | [______________] |

#### PART 2: IDI INTRODUCTION

| | |
|---|---|
| **District ID** [___] | **Interviewer initials** [___\|___\|___] |
| **IDI ID number** [___\|___\|___] | **Note-taker initials** [___\|___\|___] |
| **Date:** [___\|___] / [___\|___] / [___\|___] (day / month / year) | **Time start** [___\|___] : [___\|___]<br>**Time end** [___\|___] : [___\|___] |

**Introduction**

I am ______________________ from ______________ (interviewer)
I am ______________________ from ______________ (note-taker)

- *Hello. My name is.... and I work for IDRC (Infectious Diseases Research Collaboration).*
- *Thank you very much for meeting us today. I am part of a team who are doing research to learn more about peoples' experiences with immunization services. We would like to understand more about what you think about immunization, what has gone well when it comes to seeking and receiving immunization services and the challenges you have faced. In addition to our discussion with you today, we are talking with other mothers, fathers and primary care providers, VHTs and health care workers in this district.*
- *Today we would like you to take part in a discussion about immunization of children, your experiences seeking and receiving immunization services and what can be done to improve immunization services in your community. We have invited you to participate because you have experience with immunisation here and we hope you will tell us the real situation from your point of view.*
- *The discussion will last for about one hour. We will take notes of the ideas discussed and, if you agree, a recording will be made of the discussion in order that we record what you say accurately. When the tape is on, the light will be red. If you wish to say anything 'off the record' this is fine, please indicate to the interviewer or note taker. The audio recording will only be used by the study team: no one else will hear your voice. It will be kept for 2 years for our records and will then be erased or destroyed. We are not writing down your names here and no one will be able to identify you in any reports arising out of this research. All records of this discussion will be kept securely.*
- *Does anyone have any questions?*
- *Let us begin by setting some ground rules.*
  - ✓ Important things to emphasize
    - It is important for us to hear all sides of an issue – the positive and the negative.
    - Confidentiality is assured. "What is shared in the room stays in the room.
    - TURN OFF MOBILE PHONES OR KEEP THEM IN SILENT MODE
  - ✓ Consent

MAKERERE UNIVERSITY
SCHOOL OF PUBLIC HEALTH
APPROVED
VALID UNTIL
★ 01 AUG 2024 ★
RESEARCH & ETHICS COMMITTEE
P. O. BOX 7072, KAMPALA

#### PART 3: IDI TOPIC GUIDES (1)

**Domains, topic questions, and probes:** Use the table below to help you administer the questions during the interview.

*'Now I am going to introduce some topics one at a time about your experiences when your children were immunized, and I hope you can discuss them together.'*

| Domain | Topic and Probes |
|---|---|
| **1. Common illnesses in children** | a) Tell me about the children under your care. How many are they? How old are they? |
| | b) What illnesses have been common in children under 5 years here for the last few months? *(Make a list, don't spend too long on this question)* |
| **2. Thinking and Feeling about vaccines** | a) What have you heard about vaccines for children? *(Probe for when they heard this and the source of this information)* |
| | b) Do you think vaccines are needed for children? Why are vaccines needed for children? *(Probe for reasons as to why they think vaccines are needed or not needed for children)* |
| **3. Experiences with vaccination & social processes** | a) Where do you go to receive immunization services in this community? (*Probe for how far the place is from where they stay and how they usually get to the place where immunization services are given*) |
| | b) Tell me about the last time any of your children were vaccinated with DPT vaccine (*DPT injection is given on the left thigh)*. Why did you decide to get your child(ren) vaccinated? |
| | c) [illegible] did you consult or seek advice from before deciding to get your child vaccinated? |
| | d) What did you like about the services that you received that day? *(Probe: What was it about that experience that made you feel satisfied?)* What did you not like about the services that you received that day? |
| | e) What difference has getting your child vaccinated made now or will it make in the future? |

MAKERERE UNIVERSITY
SCHOOL OF PUBLIC HEALTH
APPROVED
VALID UNTIL
★ 01 AUG 2024 ★
RESEARCH & ETHICS COMMITTEE
P O. BOX 7072, KAMPALA

| PART 3: IDI TOPIC GUIDES (2) | |
|---|---|
| **Domains, topic questions, and probes:** Use the table below to help you administer the questions during the interview. | |
| **Domain** | **Topic and Probes** |
| **4. Practical issues & Challenges with seeking immunization services** | a) Tell me about the situations when you have failed to take your child(ren) for immunization. What were the reasons for not taking your child(ren) for immunization? *(Probe for examples and stories)* |
| | b) What challenges have you encountered in getting your child(ren) vaccinated at the recommended schedule? (*Probe for challenges related to availability of vaccines, costs, service quality, ease of access*) |
| | c) How did you overcome these challenges to ensure that your child(ren) got vaccinated? |
| **5. Suggestions for improving vaccination services** | a) What improvements would you like to see in the way immunization services are provided in this community/at your health centre? |
| | b) What are your suggestions on how the Ministry of Health can ensure that all Ugandan children get vaccinated? |

| | |
|---|---|
| **6. Closing** | We are now approaching the end of our discussion. Is there anything else you would like to add about immunization services in your community/ at your health centre that we have not talked about? |
| ✓ **Summarise**<br>✓ **Thank participant** | |

MAKERERE UNIVERSITY
SCHOOL OF PUBLIC HEALTH
APPROVED
VALID UNTIL
★ 01 AUG 2024 ★
RESEARCH & ETHICS COMMITTEE
P. O. BOX 7072, KAMPALA

| PART 3: CONTACT SUMMARY FORM (1) | |
|---|---|
| Complete this form after the interview. | |
| Study ID<br>[____\|____] | Date<br>[____\|____] / [____\|____] / [____\|____]<br>day month year |
| 1. How would you describe the atmosphere and context of the interview (*Include interview location and how this may have affected responses)*? | |
| 2. What were the main points made by the respondent during this interview? | |

MAKERERE UNIVERSITY
SCHOOL OF PUBLIC HEALTH
APPROVED
VALID UNTIL
★ 01 AUG 2024 ★
RESEARCH & ETHICS COMMITTEE
P O. BOX 7072, KAMPALA

| **PART 3: CONTACT SUMMARY FORM (2)** | |
|---|---|
| **Study ID**<br>[____ \| ____] | **Date**<br>[____ \| ____] / [____ \| ____] / [____ \| ____]<br>day month year |
| 3. What new information did you gain through this interview compared to previous interviews? | |
| 4. Was there anything surprising to you personally? Or that made you think differently? | |
| 5. What messages did you take from this interview to improve the uptake of immunization services? | |
| 6. Were there any problems with the topic guide (e.g. wording, order of topics, missing topics) you experienced in this interview? | |

MAKERERE UNIVERSITY
SCHOOL OF PUBLIC HEALTH
APPROVED
VALID UNTIL
★ 01 AUG 2024 ★
RESEARCH & ETHICS COMMITTEE
P O. BOX 7072, KAMPALA

### APPENDIX B: KEY INFORMANT INTERVIEW TOPIC GUIDE
#### VHTs/Community Leaders

| Study ID | Date |
|---|---|
| [____\|____] | [____\|____] / [____\|____] / [____\|____]<br>day month year |

**Position:**
1 = Village Health Team member
2 = Religious Leader
3 = Opinion Leader
4 = Political Leader
5 = Local Chairman
6 = Other________________________

[____\|____
]

#### DEMOGRAPHIC INFORMATION

| | | | |
|---|---|---|---|
| **1. Age** | Years | [____\|____] | **5. Highest level of education or qualification achieved** |
| **2. Gender** | 1 = Male<br>2 = Female | [____] | 0 = None<br>1 = Primary (P1 — P7)<br>2 = Secondary (S1 — S6)<br>3 = Certificate<br>77 = Other________________<br>4 = Diploma<br>5 = Bachelor's degree<br>88 = Don't know<br>99 = Refused to answer |
| **3. Originally from this area?** | 1 = Yes<br>2 = No | [____] | |
| **4. Number of years worked in this job/role** | | [____\|____] | [____\|____] |

#### PART 1: INTRODUCTION

Conduct the interview according to the directions below and record information as indicated.

**Introduction to interview**

*"Hello, my name is .......... I work with IDRC (Infectious Diseases Research Collaboration). I am interested in asking you a few questions about immunization activities, services or other contextual factors occurring in this district. A note-taker will be writing down what you say for our record and we will record the interview using a digital recorder; these notes will be kept securely and your name will not be used anywhere. Your answers will be looked at together with those of many other VHTs, local leaders and health care providers involved in the planning and delivery of immunization services, and you will not be identifiable in any reports that are published.*

*It is very important for us to hear your views and experiences because your knowledge and experience can give insight to our study. We hope you will have time to spend with us now to complete this interview. The interview will take about 45-60 minutes; if you prefer we can reschedule the interview for tomorrow or another day of your convenience.*

*Do you have any questions? Do you agree to continue before we start?*

*Now we request that we all switch off our mobile phones or keep them in silent mode so that we are not distracted."*

MAKERERE UNIVERSITY
SCHOOL OF PUBLIC HEALTH
APPROVED
VALID UNTIL
★ 01 AUG 2024 ★
RESEARCH & ETHICS COMMITTEE
P O. BOX 7072, KAMPALA

#### PART 3: KII TOPIC GUIDES (1)

**Domains, topic questions, and probes:** Use the table below to help you administer the questions during the interview.

*'Now I am going to introduce some topics one at a time about immunization services in this area, and I hope you can discuss them together.'*

| Domain | Topic and Probes |
|---|---|
| 1. Description of Job | a) Can you briefly describe your roles and responsibilities in your job? |
| | b) What specific role do you play in immunisation-related programmes? |
| 2. Thinking and Feeling about vaccines | a) What have you heard about vaccines for children? *(Probe for when they heard this and the source of this information)* |
| | b) Do you think vaccines are needed for children? Why are vaccines needed for children? *(Probe for reasons as to why they think vaccines are needed or not needed for children)* |
| | c) Drawing on your observations and experiences, what has been the response of people in this area to immunization services for children? (Probe: How have immunization services for children been received by people in this community?) |
| | d) What do people in this area like about getting their children immunized? What do they see as the benefits of getting their children immunized? |
| | e) What concerns or worries do people in this area have when it comes to their children getting immunized? |

MAKERERE UNIVERSITY
SCHOOL OF PUBLIC HEALTH
APPROVED
VALID UNTIL
★ 01 AUG 20[illegible] ★
RESEARCH & ETHICS COMMITTEE
P O. BOX 7072, KAMPALA

| PART 3: IDI TOPIC GUIDES (2) | |
|---|---|
| **Domains, topic questions, and probes:** Use the table below to help you administer the questions during the interview. | |
| Domain | **Topic and Probes** |
| **3. Practical issues & Challenges with seeking immunization services** | a) Where do people go to receive immunization services in this community? (*Probe for how accessible these places are for the people targeted for immunization services in the local area. What is the preferred place (health facility or outreaches) for immunization in this community?*) |
| | b) Overall, what challenges do people in this area encounter in getting their child(ren) vaccinated at the recommended schedule? (*Probe for challenges related to availability of vaccines, costs, service quality, ease of access*) |
| | c) Are you aware of communities who have a significant number of children who have never been immunised/are partially vaccinated or under immunised communities in your area? Please tell me about the zero dose, under immunised and missed communities. (Probe for who they are) |
| | d) What are the reasons for children/communities not being immunized or not completing all the doses e.g., children receiving DPT1 (first dose) but do not return for DPT3 (third dose) and some communities being missed? |
| | e) What strategies or interventions have been undertaken in this community to address the challenges listed above and ensure that all children in your community receive vaccines? (Probe for whether these strategies have worked or not and why?) |
| | f) How can these strategies be improved to better reach the zero dose, under immunised and missed communities with vaccines? |

MAKERERE UNIVERSITY
SCHOOL OF PUBLIC HEALTH
APPROVED
VALID UNTIL
★ 01 AUG 2024 ★
RESEARCH & ETHICS COMMITTEE
P O. BOX 7072, KAMPALA

| | |
|---|---|
| 4. Social mobilization & sensitization of communities on vaccination | a) What activities are undertaken to prepare communities for immunisation programmes before they are rolled out? (*Probe for social mobilization and community sensitization activities, what they have involved, who has been involved, channels used*) |
| | b) How helpful have these activities been in getting people to agree to take up immunisation services for their children? (*Probe for what has worked well and what the challenges have been)* |
| | c) What is the best way to inform communities about vaccines and mobilise them for vaccination programs? (Probe for who should be involved, the best channels to be used) |
| 5. Suggestions for improving vaccination services | a) What improvements would you like to see in the way immunization services are provided in this community? |
| | b) What are some ways in which immunization programs can better reach all children in your community with vaccines? |

| | |
|---|---|
| 6. Closing | We are now approaching the end of our discussion. Is there anything else you would like to add about immunization services in your community or non-and under immunization in Uganda that we have not talked about? |
| ✓ **Summarise**<br>✓ **Thank participant** | |

MAKERERE UNIVERSITY
SCHOOL OF PUBLIC HEALTH
APPROVED
VALID UNTIL
★ 01 AUG 2024 ★
RESEARCH & ETHICS COMMITTEE
P O. BOX 7072, KAMPALA

| PART 3: CONTACT SUMMARY FORM (1) | |
|---|---|
| Complete this form after the interview. | |
| Study ID<br>[____\|____] | Date<br>[____\|____] / [____\|____] / [____\|____]<br>day month year |
| 1. How would you describe the atmosphere and context of the interview (*Include interview location and how this may have affected responses)*? | |
| 2. What were the main points made by the respondent during this interview? | |

MAKERERE UNIVERSITY
SCHOOL OF PUBLIC HEALTH
APPROVED
VALID UNTIL
★ 01 AUG 2024 ★
RESEARCH & ETHICS COMMITTEE
P O. BOX 7072, KAMPALA

MAKERERE UNIVERSITY
SCHOOL OF PUBLIC HEALTH
APPROVED
VALID UNTIL
★ 01 AUG 2024 ★
RESEARCH & ETHICS COMMITTEE
P.O. BOX 7072, KAMPALA

#### PART 3: CONTACT SUMMARY FORM (2)

| Study ID | Date |
|---|---|
| [____\|____] | [____\|____] / [____\|____] / [____\|____] <br> day month year |

3. What new information did you gain through this interview compared to previous interviews?

4. Was there anything surprising to you personally? Or that made you think differently?

5. What messages did you take from this interview to improve the uptake of immunization services?

6. Were there any problems with the topic guide (e.g. wording, order of topics, missing topics) you experienced in this interview?
